# Increased Hindbrain Motion in Chiari Malformation I Patients Measured Through 3D Amplified MRI (3D aMRI)

**DOI:** 10.1101/2022.10.25.22281481

**Authors:** Javid Abderezaei, Aymeric Pionteck, Ya-Chen Chuang, Alejandro Carrasquilla, Gizem Bilgili, Tse An Lu, Itamar Terem, Miriam Scadeng, Patrick Fillingham, Peter Morgenstern, Michael Levitt, Richard G. Ellenbogen, Yang Yang, Samantha J. Holdsworth, Raj Shrivastava, Mehmet Kurt

## Abstract

Chiari Malformation type 1 (CM-I) is a neurological disorder characterized by morphological defects such as excessive cerebral tonsils herniation and vast associated symptomatology. Given that these structural defects cannot explain the underlying symptomatology, and might result in misdiagnosis, in this work, we studied the brain’s intrinsic motion to better understand the mechanisms of CM-I. We acquired 3D cine MRI of 14 healthy and 14 CM-I subjects and used 3D amplified MRI (3D aMRI) to visualize and measure the brain’s intrinsic motion during the cardiac cycle. We observed that the regional brain motion in CM-I was significantly higher than the healthy subjects, with anterior-posterior (AP) and superior-inferior (SI) displacements in cerebral tonsils and medulla having the highest differences between the healthy and CM-I (∽45% ∽73% increased motion in the CM-I group). The motion of the cerebellum, and brainstem in AP directions (∽42% and ∽31% increased motion in the CM-I group, respectively), followed by the motion of the cerebral tonsils and medulla in medial-lateral (ML) directions were other significant differences found between the two groups (∽16% increased motion in the CM-I group). Additionally, for the CM-I subjects, we measured morphological parameters including the tonsil herniation, ratio of neural tissue in the foramen magnum, and 4^*th*^ ventricle volume. We then used the morphometrics and brain’s intrinsic motion to analyze the symptomatology of the CM-I patients and their surgical outcomes. Interestingly, we found the ratio of neural tissue in the foramen to be directly correlated with the SI motion of the tonsils (*r* = 0.58). We also found the tonsil herniation to be directly correlated with the AP motion of the tonsils (*r* = 0.61), and AP and ML motions of the medulla (*r* = 0.66, and *r* = 0.57). Additionally, we found the ML motion of the tonsils to be the only indicator of the surgical outcome (*AUC* = 0.95), in which subjects with higher motion had an improved outcome. Although we did not observe a significant correlation between the brains motion and morphometrics on the CM-I symptoms due to our small sample size, illustrative cases increase our hope for the development of a future tool based on the brain biomechanics.

## 1. Introduction

Chiari Malformation Type I (CM-I) is a common and often debilitating craniocervical junction neurological disease which is diagnosed in about 1% of the adults and 3.6% of the children population who receive MRI scans [1–4]. CM-I is characterized by structural defects in the cerebellum; more specifically, at least a 3-5 mm downward herniation of the cerebellar tonsils through the foramen magnum [2, 5]. It is associated with a vast symptomatology, such as recurrent headaches, nausea, muscle weakness, sleep disorders, and in the more extreme cases, paralysis [1, 6, 7]. These symptoms may begin to appear early in the development, or only during the adult life [8–10]. This is particularly important in the adult population, since early diagnosis of CM-I and subsequent surgical treatment can lead to a dramatic improvement in clinical outcomes [8]. Currently, the diagnosis of CM-I is based on an assessment of the patient’s neurological history combined with an MRI or CT examination. However, the lack of a uniform and clear symptomatology among the subjects is so pronounced that an estimated 30% of the patients affected do not show symptoms significant enough to lead to a diagnosis [6, 11]. This inconsistency is so striking that, in a recent study, about 2 times more asymptomatic cases than the symptomatic ones were found in the CM-I subjects diagnosed by measuring the herniation of the cerebral tonsils [1]. Another study reported that about 14% of the patients with an abnormal cerebral tonsil do not have any symptoms [2, 6]. This lack of clear and uniform symptomatology could lead to misdiagnosis and clinical complications. At the same time, it is equally important to determine the patient to be operated on, as the surgery can carry morbidity [12]. In a recent study, it was shown that within 30 days of the surgery on CM-I patients, approximately 14.3% experienced surgical complications, and about 4.4% of them experienced other medical complications [12]. These numbers rose to 18.7% and 5%, 90 days after the surgical operations for these CM-I patients [12]. Others have found that about 25% the CM-I patients either did not experience improvement postsurgery or even got worse [13]. They also reported that only about 40% of the patients had significant improvement post-surgery [13]. Another important factor to consider is the additional healthcare economic cost of surgery for misdiagnosed patients, further emphasizing the need to better understand CM-I and the underlying etiology. To date, there is still controversy regarding the etiology of the malformation and, more importantly, in which cases treatment is necessary [14–17]. Since so many patients have malformation but display no symptoms, morphological defects alone are not sufficient to induce symptoms. Therefore, there must be a more complex neurophysiological mechanism involved in the disease etiology than just a simple malformation. One such mechanism might be related to the intrinsic brain motion during the cardiac cycle.

The brain could be considered as a complex mechanical system, in which the tissue, blood flow and CSF circulation are linked, causing an intrinsic pulsatile brain movement that could be investigated to better understand brain diseases with obstructive disorders. CM-I is one such disease, in which morphological changes in the cerebellum can lead to obstruction of the subarachnoid space near the foramen magnum [18, 19]. The obstruction of the CSF flow through this region could ultimately affect the brain’s intrinsic motion during the cardiac cycle [18, 19]. But, until now, the effect of physiological flow such as the blood flow and CSF circulation on the intrinsic brain motion has proven difficult to analyze. The main reason for this difficulty is that the intrinsic motion of the brain is extremely small (in the order of 100 *µm* [20]), and therefore, very difficult to measure with conventional imaging tools. The most prominent method to track the brain motion is through magnetic resonance elastography (MRE) [21– 25] in which an external actuator [26] is used to cause shear wave propagation into the brain tissue and then the resultant tissue displacement field is captured by using Phase-contrast MRI (PC-MRI) [22–24, 27]. To capture the intrinsic brain motion without an external actuator however, previous medical imaging studies used cine PC-MRI [28], or displacement encoded imaging with stimulated echoes (DENSE) MRI [20, 29, 30]. However, these studies have been limited by imaging resolution (since the displacements studied are very small), or have only been able to analyze the brain motion in a single sagittal slice (without accounting for out-of-plane motion [29]). AMRI is another imaging technique that has also been used to analyze the brain’s intrinsic motion [31, 32]. AMRI has not only been used for visualizing the CM-I brain motion [32], but also, its modified version called amplified flow imaging (aFlow) has also been used for the analysis of the intracranial aneurysm wall motion [33, 34]. More specifically for CM-I, the 2 directional (2D) version of aMRI was applied on one patient and an increased caudal midbrain tissue motion and a downward motion at the level of the brainstem and craniocervical junction was observed in the CM-I subject as compared to the healthy ones [32]. AMRI, however, was only able to track the intrinsic brain motion in 2D [31, 32]. More recently, our group extended the 2D aMRI method to 3D, which allowed visualization and quantification of the brain’s intrinsic motion in 3 directions [35, 36]. In these works, superior-inferior motion near the pons and the 4^*th*^ ventricle were found to be the highest in healthy subjects [35, 36].

In the current work, we applied the 3D aMRI algorithm to a cohort of healthy and CM-I patients. We then quantified the amplified brain motion in 3D using an image registration algorithm. After performing a semiautomatic segmentation of the brain, we quantified the motion in ROIs including the cerebellum, cerebral ton-sils, brainstem, and medulla and compared the displacements between the healthy and the CM-I group. Next, we measured morphological parameters including the cerebral tonsil herniation, the ratio of neural tissue in the foramen magnum, and the 4^*th*^ ventricle volume for the CM-I subjects. Finally, we analyzed the intrinsic brain motion and morphometrics in the CM-I group and calculated the correlations between these parameters and the existence of symptoms and the surgical outcomes.

## 2. Methods

### 2.1. Human subjects

With IRB approval and informed consent (IRB-18-01303), scans were performed on 14 healthy adult volunteers (7 females and 7 males between 24 and 60 years of age (41.0 ± 11.4)) and 14 CM-I patients (12 females and 2 males between 24 and 68 years of age (39.0 ± 13.5)) using a 3T (Skyra, Siemens Healthcare AG, Germany) MR imaging system with a 32-channel head coil at Icahn School of Medicine at Mount Sinai. In addition to acquiring the MR images of the subjects, common clinical symptoms of the CM-I subjects were recorded pre and post decompression surgery (Table. 1). 9 of the 14 CM-I subjects underwent a posterior cranial fossa decompression surgery, 4 of which had an improved outcome, whereas, the other 5 patients either did not get better or only had minor improvements post-surgery. These symptoms and the surgical outcomes were recorded by neurosurgeons at Icahn School of Medicine at Mount Sinai.

**Table 1:** Clinical symptoms of the CM-I subjects.

| Symptoms | Number of subjects |
| --- | --- |
| Headache | 13 |
| Dizziness | 5 |
| Back, neck, ear pain | 8 |
| Gait disturbance | 11 |
| Visual symptoms | 6 |
| Nausea and vomit | 2 |
| Tinnitus and vertigo | 3 |
| Weakness | 3 |

### 2.2. Imaging protocols

A cardiac-gated bSSFP sequence [37] was used to acquire 3D volumetric data of brain’s intrinsic motion. Heartbeat and MRI data acquisition were synchronized using a pulse oximeter to generate a 3D cine sequence of the brain over one cardiac cycle. Whole-brain 3D cine MRIs were acquired using the following imaging parameters: acquisition matrix = 240 × 173, flip angle = 26°, field-of-view = 216 × 240 mm^2^, repetition time/echo time = 36.6/1.6 ms, pixel bandwidth = ± 990 kHz, 16-23 cardiac phases (temporal resolution of 40-60 ms, depending on the subject’s heart rate), and 80 slices with a spatial resolution of 1 × 1 × 1.2 mm^3^ in anterior-posterior (AP), superior-inferior (SI) and medial-lateral (ML) directions, respectively.

### 2.3. 3D aMRI algorithm

The 3D aMRI method [35, 36] is an extension of the 2D aMRI algorithm [31, 32], allowing for the visualization and quantification of sub-voxel level motion in 3 directions. In the first step, a complex steerable pyramid in the spherical domain is applied to the input data, which in our study, is 3D cine MRI datasets of healthy and CM-I subjects. By using the 3D steerable pyramid the volumetric data is decomposed at different angles and scales. In the next step, the temporal phase differences of the decomposed data is calculated and a temporal band-pass filter is applied to isolate the motion at the selected frequency bands. To reduce the noise and artifacts, a Gaussian spatial smoothing filter can be optionally added to the next step. The processed phase differences at each angle and scale are then amplified using an amplification factor *α* and then added back to the original reference phase. Finally, the 3D steerable pyramid is collapsed and the decomposed data is reconstructed which results in a 3D amplified data. For more information about the 3D aMRI algorithm please see [35, 36].

### 2.4. 3D aMRI analysis of CM-I and healthy in vivo data

In this step, the 3D cine MRI of the healthy and CM-I subjects were amplified using the 3D aMRI algorithm. We chose *α* = 8 and frequency band *f* ∈ [0 4] Hz as the amplification parameters [35]. These parameters were selected based on our previous results for healthy subjects [35]. Next, the Demons registration algorithm [38] was applied to the amplified data, and the 3D brain displacements of these subjects were calculated. Here, we analyzed the displacements of 4 brain regions including cerebellum, cerebral tonsils, brainstem, and medulla, since previous studies have reported abnormal motion of these brain structures in CM-I patients [29, 39].

### 2.5. Semi-automatic brain segmentation technique

In order to create masks and measure the displacements of these brain structures, a semi-automated brain segmentation method was used (Fig. 1). To do so, the T1-weighted MRI of each subject was initially processed with FreeSurfer which allowed extracting masks for the cerebellum, brainstem and medulla [40–42]. The cerebral tonsils of each subject was then manually segmented. The automatically generated masks of each subject were then manually modified in order to improve their accuracy. The general pipeline of the semiautomatic image segmentation can be seen in Fig. 1. An automatic intensity based image registration method was next used in MATLAB (MathWorks, Natick, MA, version R2020b) to find the transformation matrix between the T1-weighted MRI of each subject and its corresponding 3D cine MRI. The calculated transformation matrix was then applied to the created masks for the cerebellum, cerebral tonsils, brainstem, and medulla, to register the T1-weighted MRI based masks to their corresponding 3D cine MRI. Each mask for the 3D cine MRI data was then manually inspected in order to confirm its accuracy.

**Figure 1:**
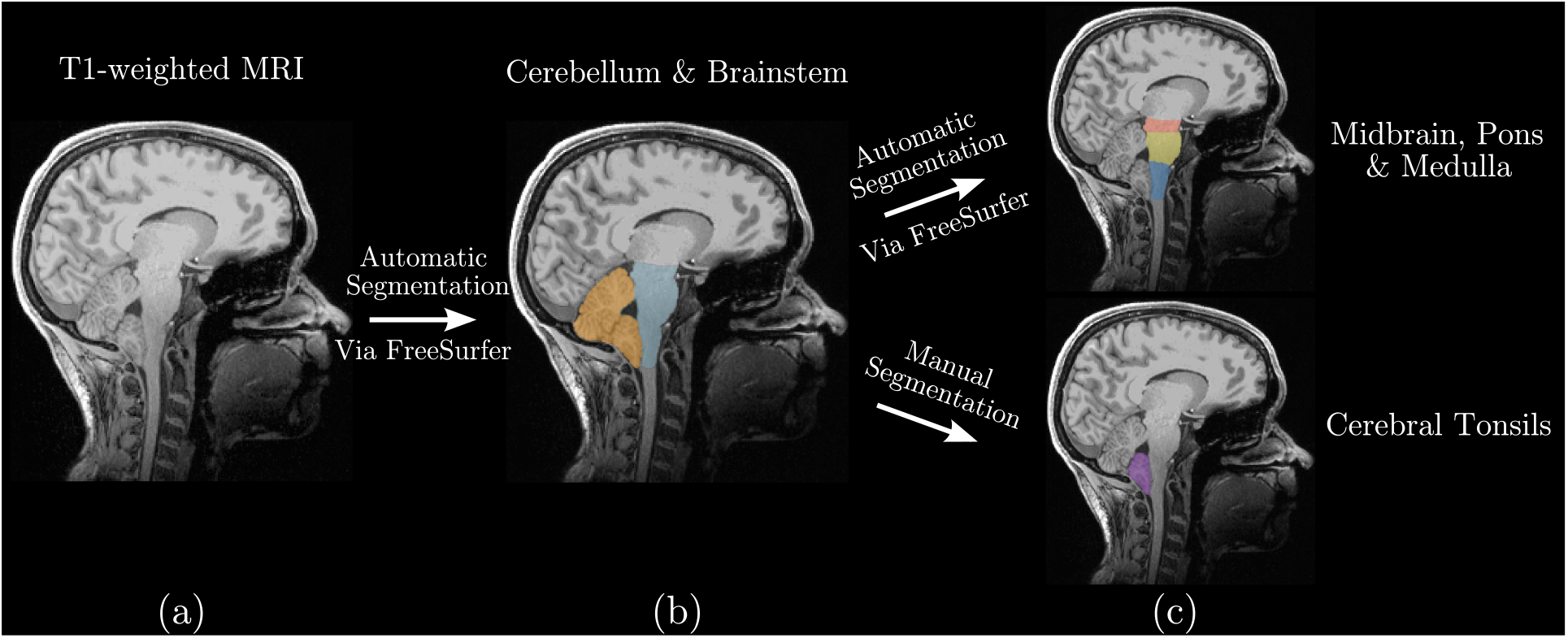
Schematics of the semi-automatic segmentation of the cerebellum, cerebral tonsils, brainstem, and medulla. a) T1-weighted MRI of a CM-I patient. b) Cerebellum and brainstem were automatically segmented via FreeSurfer (orange and blue, respectively). c) Medulla was segmented automatically via FreeSurfer (blue), whereas, the cerebral tonsil was manually segmented (purple).

### 2.6. Displacement measurement

In this step, we calculated the maximum displacement in AP, SI, and ML directions, as well as their magnitude for the 4 brain segmentation (*i*.*e*. cerebellum, cerebral tonsils, brainstem and medulla). Below, we calculated the displacement magnitude |U(x, y, z, t) | of each voxel at coordinate x, y, z, and time t as:

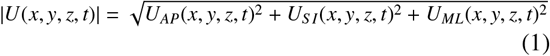

where *U*_*AP*_(*x, y, z, t*), *U*_*S I*_(*x, y, z, t*), and *U*_*ML*_(*x, y, z, t*) are the displacements in AP, SI, and ML directions. Next, we calculated the maximum temporal displacement of each voxel as *U*_*AP*,*Max*_, *U*_*S I*,*Max*_, *U*_*ML*,*Max*_ and *U*_*Mag*,*Max*_ for the three directions and magnitude, respectively. Finally, we calculated the 90^*th*^ percentile of the maximum temporal displacements of the voxels within each brain segmentation for different directions 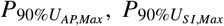, and 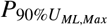 and maximum magnitude 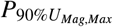.

### 2.7. Morphometric analysis

In addition to the evaluation of the intrinsic brain motion, we measured various morphological parameters for each of the CM-I patients. These parameters include: Cerebral tonsil herniation, the area of foramen magnum, the ratio of neural brain tissue that occupies the foramen magnum, and the 4^*th*^ ventricle volume.

To measure the cerebral tonsil herniation, we first centered the images with respect to the coronal and axial planes. The mid-sagittal slice was then selected and the foramen magnum was indicated by connecting a line from basion to the opisthion (McRae’s line). Cerebral tonsil herniation was measured by calculating the perpendicular distance from the McRae’s line to the end of the tonsil.

We then selected the axial plane at foramen magnum and measured the surface areas of cervicomedullary junction *A*_*C*_, the tonsil section *A*_*T*_, and the total surface area of the foramen magnum (*A*_*FM*_). Finally, we calculated the ratio of neural tissue in the foramen magnum by using (*A*_*T*_ + *A*_*C*_)*/A*_*FM*_. These measurements were performed by 3 of the authors using FreeSurfer and Horos (GNU Lesser General Public License, Version 3 (LGPL-3.0)), and the results were averaged. Finally, the 4^*th*^ ventricle segmentation was initially estimated by the FreeSurfer software and was then manually corrected by one of the authors and the resulting volume size was calculated.

### 2.8. Statistical analysis

Our statistical analysis can be divided into two parts. In the first part, we compared the healthy brain motion and the CM-I subjects. In the second part, we only considered the CM-I subjects and analyzed the correlations between the brain motion, morphometrics, symptomatology and surgical outcomes.

To statistically compare the CM-I and healthy brain motion, t-tests and Wilcoxon rank-sum tests were performed between 3 directional displacements (*i*.*e*.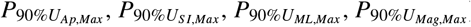) at each of the selected ROIs including the cerebellum, cerebral tonsils, brainstem, and medulla.

In the following steps, we only used the CM-I dataset and analyzed the correlations between the brain motion, morphometrics, symptomatology and surgical outcomes. First, we calculated the Pearson correlation values (*r*) between the morphometrics including the cerebral tonsil herniation, the ratio of neural tissue in the foramen magnum ((*A*_*T*_ + *A*_*C*_)*/A*_*FM*_), and the 4^*th*^ ventricle volume and each of the 3 directional brain displacements in the selected ROIs. In the next step, we used the ROI brain displacements and morphometrics and performed a comparison between the subjects with and without the symptoms. Next, we used the ROI brain displacements and morphometrics to perform a similar analysis for the subjects with and without improved surgical outcome. Finally, for the above symptomatology and surgical outcome analysis, we also used logistic regression and calculated accuracy scores by using the Area Under the ROC Curve (AUC) [43–45].

For all the above statistical analysis, we considered the differences between the two groups to be statistically different if *p <* 0.05. Additionally, since we compared a large number of parameters in this study (4 parameters in 4 different brain regions), we also reported the family-wise error corrected of the differences by adjusting the statistical significance to *p <* 0.0031 (0.05*/*16) by using the Bonferroni correction [46].

## 3. Results

### 3.1 Comparison of the brain displacement between the healthy and CM-I subjects

In the first step, we amplified the brain motion in the 14 CM-I and 14 healthy controls and calculated the displacement fields of each subject. We then masked the brain ROIs including the cerebellum, cerebral tonsils, brainstem, and medulla and compared the displacements between the two groups. In cerebellum, the 90^*th*^ percentile of the AP maximum displacement 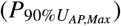 and magnitude displacement 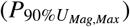 were on average about 41.96% and 22.34% higher in CM-I as compared to the healthy subjects (*p <* 0.0001 and *p <* 0.001, respectively; Table. 2, Fig. 2). In this brain region, after adjusting for the family-wise error (*p <* 0.003 = 0.05*/*16) both 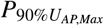 and 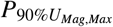 were still statistically higher in CM-I as compared to the healthy group (Table. 2). In cerebral tonsils, the magnitude, AP, SI and ML displacements were significantly higher in CM-I as compared to the healthy subjects (Table. 2). For magnitude, AP, and SI, we found *p <* 0.0001; whereas, for ML, we found *p <* 0.01 (Table. 2). Here, 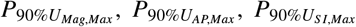, and 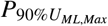 were about 49.53%, 73.38%, 43.55%, and 16.18% higher in the CM-I subjects versus the healthy controls, respectively (Table. 2, and Fig. 2). Additionally, it should be mentioned that after correcting for the family-wise error, 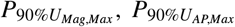, and 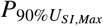 were still significantly higher in CM-I as compared to the healthy subjects (*p <* 0.003, Table. 2). We had similar findings in the displacements of the medulla, with significantly higher magnitude, SI, AP, and ML in CM-I as compared to the healthy subjects (no statistical differences in ML direction after adjusting for family-wise error; Table. 2). Here, while we found *p <* 0.0001 for 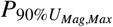, and 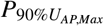, we found *p <* 0.001 for 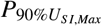, and *p <* 0.01 for 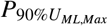, respectively (Table. 2). For Medulla, we found approximately 40.73%, 56.88%, 37.37%, and 16.87% larger 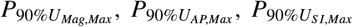, and 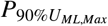 in the CM-I group as compared to their healthy controls (Table. 2, Fig. 2). In brainstem, we found 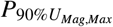, and 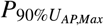 to be about 13.20%, and 31.04% larger in CM-I as compared to the healthy controls, respectively (Table. 2, Fig. 2). Here, we found *p <* 0.05 and *p <* 0.01 for 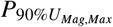, and 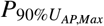, respectively. However, after adjusting the family-wise error, we only had statistical differences in 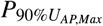 between the two groups (*p <* 0.003, Table. 2).

**Table 2:**
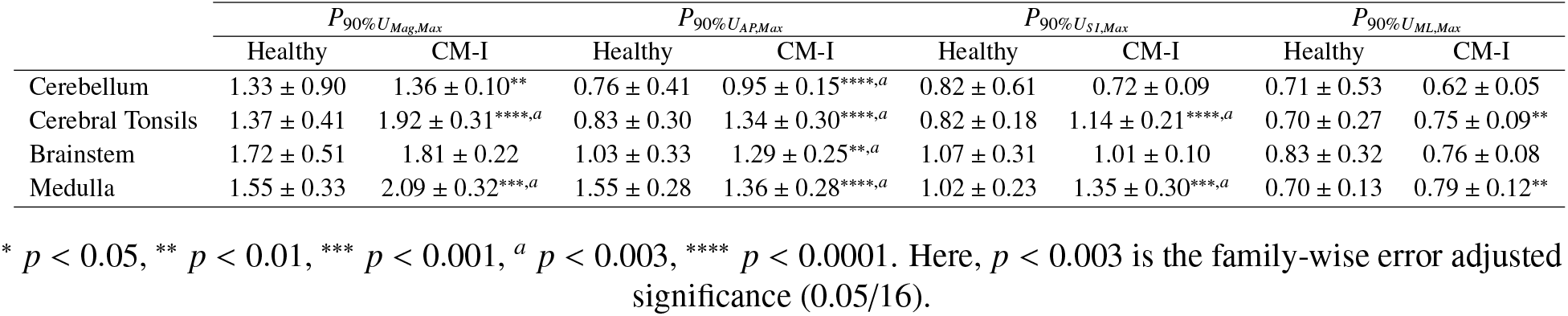
Comparison of the 90^*th*^ percentile of the minimum peak to maximum peak displacement in different directions between CM-I and healthy controls. All the units are in pixels.

**Figure 2:**
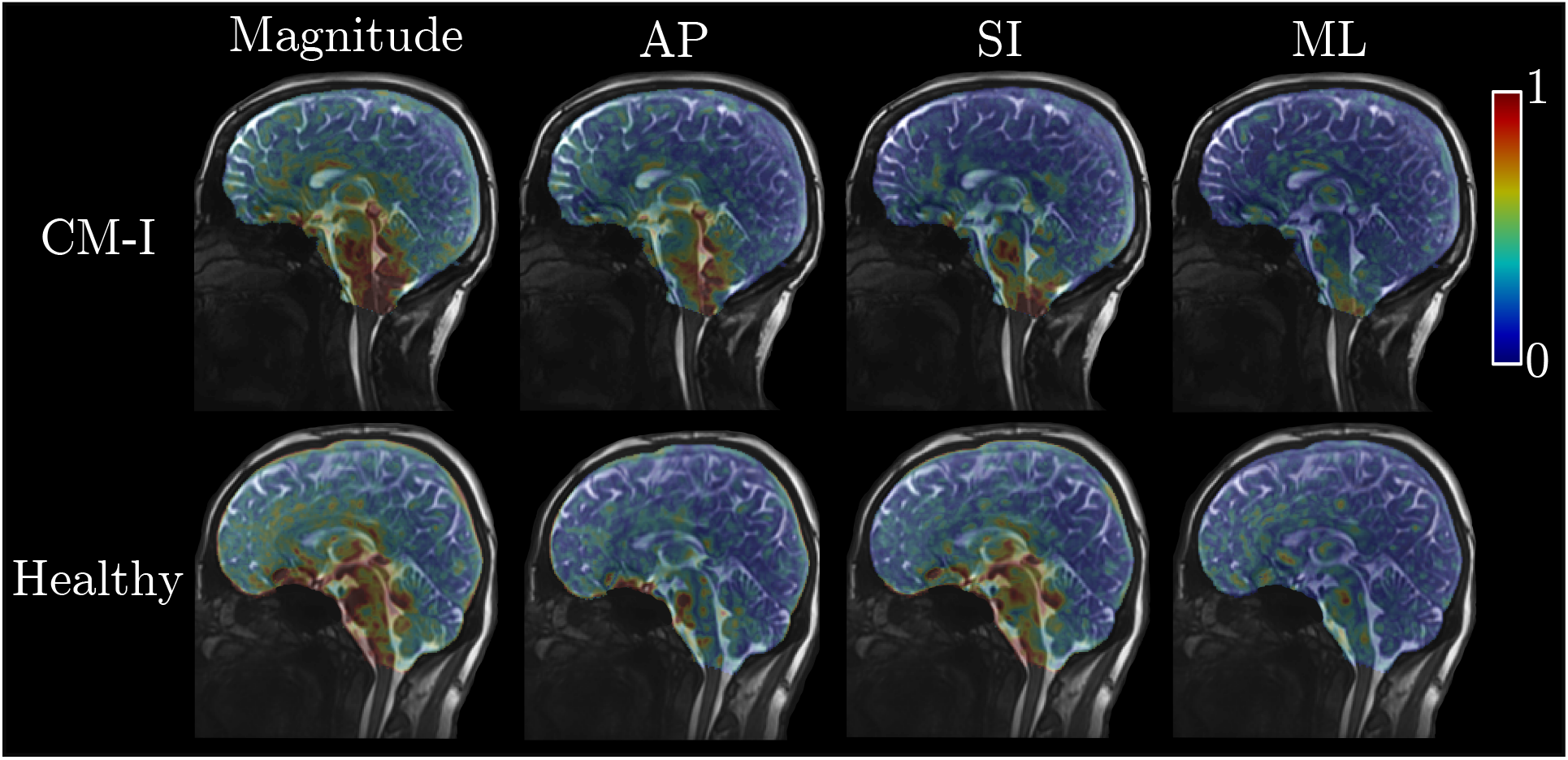
Maximum displacement of the brain in healthy and CM-I subjects during the cardiac cycle. We observed that the motion in CM-I subjects is significantly elevated in the cerebellum, brainstem cerebral tonsils and medulla. While in the cerebellum, the AP and magnitude displacement was significantly larger in CM-I, in cerebral tonsils and medulla, the magnitude, AP, SI, and ML were larger in CM-I as compared to the healthy subjects. We also found increased AP motion in the brainstem. Here, brain displacements of a CM-I and healthy female subjects in their 20s are shown as examples. *AP: anterior posterior, SI: Superior inferior, ML: medial lateral

### 3.2 Analysis of the brain motion, morphometrics, symptomatology and surgical outcomes in CM-I patients

In the next step, for the CM-I subjects we analyzed the correlations between the morphological parameters and the intrinsic brain motion at each ROI (Fig. 3, and Fig. 4). Considering the family-wise error corrected p value of *p <* 0.0031 (0.05*/*16) for significance, we did not find any of the morphological parameters to be significantly correlated with the ROI displacements. More specifically, we found Pearson correlation *r* = 0.58 between the ratio of neural tissue in the foramen magnum and the SI motion of the tonsils (*p* = 0.02, Fig. 4(a)). Additionally, we found *r* = 0.66, *r* = 0.57, and *r* = 0.61 between the tonsil herniation length and the AP, and ML motion in medulla and AP motion in tonsils, respectively (*p* = 0.01, *p* = 0.032, and *p* = 0.02, respectively; Fig. 4).

**Figure 3:**
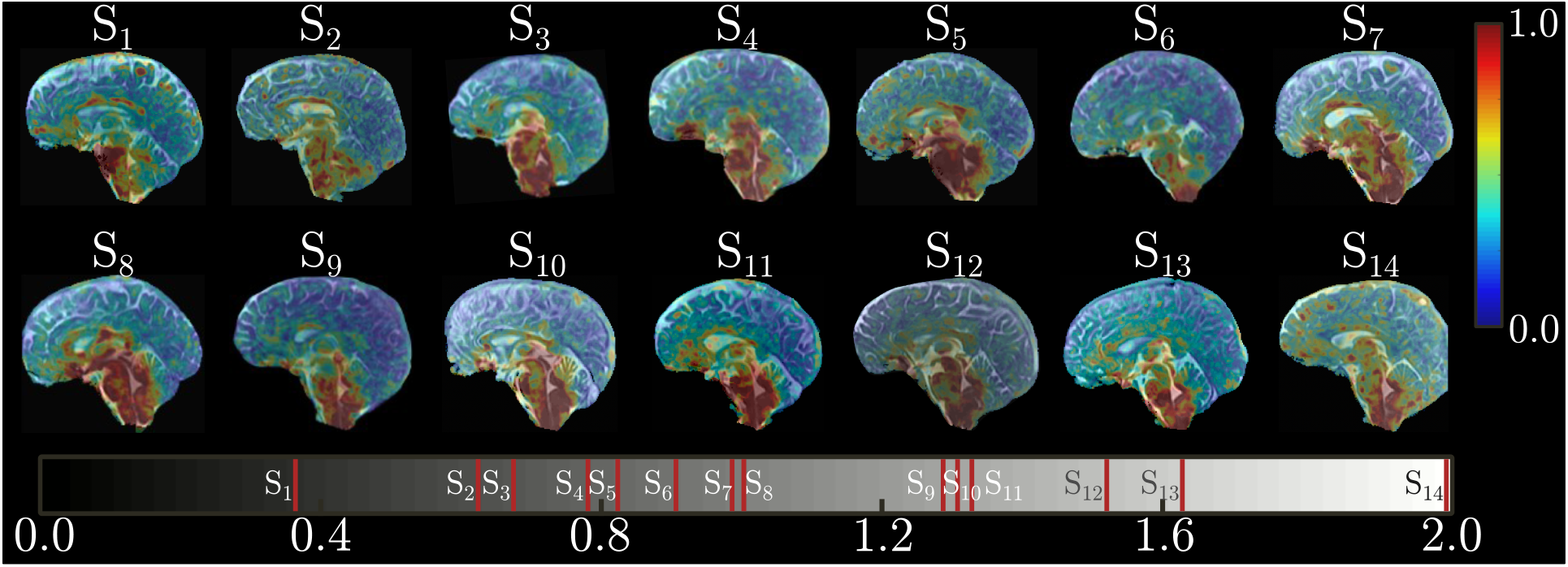
Maximum magnitude of the brain’s displacement in 14 CM-I patients with different cerebral tonsil lengths. The color bar on the right side of the figure shows the brain displacement. The gray color bar at the bottom shows the cerebral tonsil length for each subject (S). Colorbar has been limited to maximum of 1 pixel displacement to demonstrate the distribution of the displacements.

**Figure 4:**
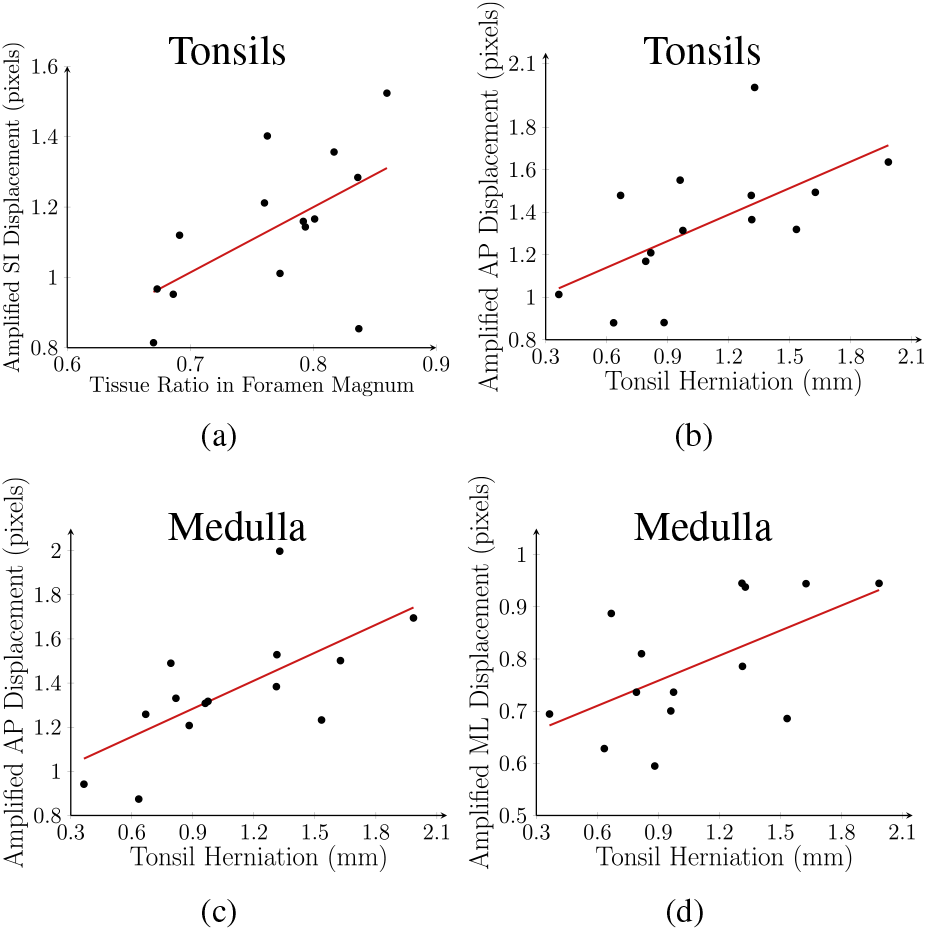
Regression analysis of the brain morphometrics and displacement of tonsils and medulla. a) A linear correlation was found between the neural tissue ratio and the amplified displacement of the tonsils in SI direction (*r* = 0.58, *p* = 0.02). b) A linear correlation was found between the cerebral tonsils herniation and the amplified displacement of the tonsils in AP direction (*r* = 0.61, *p* = 0.02), c) and medulla in AP (*r* = 0.66, *p* = 0.01), d) and ML directions (*r* = 0.57, *p* = 0.032). In the above analysis, 1 pixel = 1 mm.

Finally, we analyzed the surgical outcomes and symptomatology of the CM-I subjects by using the morphometrics and the brain’s intrinsic motion at the selected ROIs. Even though, none of the parameters was significant for the surgical outcomes using the familywise corrected p value, we found the ML displacement of tonsils to be the best indicator for pre-surgical decision making (*p* = 0.03 and AUC = 0.95). Using the 9 subjects who underwent decompression surgeries, we found that the subjects with the higher ML motion of the tonsils had improvements post-surgery, whereas, the subjects with smaller ML motion, either did not get better or had minor improvements post-surgery. More specifically, the increased ML motion of tonsils was about 11.50% higher in the CM-I subjects with improved outcome post-surgery. For the symptomatology analysis using these parameters, we did not find any statistical differences between the subjects with and without any of the 8 symptoms.

## 4. Discussion

In this study, we used 3D aMRI to quantify regional cardiac induced brain motion in CM-I and healthy subjects. We segmented regions of interests including the cerebellum, cerebral tonsils, brainstem and medulla, and analyzed their motion in 3 directions. Additionally, we used these brain displacements and various morphological parameters including the tonsil herniation, the ratio of neural tissue in the foramen magnum ((*A*_*T*_ + *A*_*C*_)*/A*_*FM*_), and the 4^*th*^ ventricle volume to analyze the CM-I subject’s symptomatology and surgical outcomes.

We observed that the regional brain motion of the CM-I as compared to the healthy controls were significantly larger in the AP direction of the cerebellum and brainstem. In cerebral tonsils and medulla, we observed this increased regional motion in all of the 3 directions (AP, SI, and ML). The reason for this increased motion in the CM-I subjects as compared to the healthy ones could be the abnormal geometry of the cerebellum which could cause increased resistance in the passage of the CSF flow through the foramen magnum [18]. What further confirms this for the CM-I subjects is the existence of direct correlations between the tonsil herniation and its AP displacement (*r* = 0.58, *p* = 0.02) as well as the direct correlations between the neural tissue crowding in the foramen magnum and the SI displacement of the tonsils (*r* = 0.61, *p* = 0.02). In the CM-I patients, the downward herniation of the cerebral tonsils cause a partial blockage of the subarachnoid space near the foramen magnum [18, 19, 47]. This partial blockage results in the obstruction of the CSF circulation at foramen magnum, which can potentially cause increased resistance and abnormal CSF flow velocities in that region [18]. This increased resistance could potentially decrease the CSF flow in that region, however, the driving force due to the arterial pressure is larger than the intracranial pressure which can ultimately result in the same volume of CSF to pass through even with the obstruction [18]. This abnormal CSF flow in the region could, therefore, be responsible for the increased motion in the AP and SI directions of the cerebral tonsils and medulla. Our findings regarding the brain motion of the CM-I subjects are in line with others [29, 48–50]. Increased AP motion near the fastigium of the 4^*th*^ ventricle as well as increased AP, and SI motion in the cerebral tonsil has been reported by Leung *et al*. [48]. Here, the maximum AP, and SI motion in the cerebral tonsils were approximately 136% and 163% higher in CM-I as compared to the healthy, respectively [48]. An increased displacement of about 64%-208% has been reported in the cerebellum and brainstem of the CM-I subjects [29]. About 18% increase in the mean of spinal cord displacement has also been reported by [51]. Some of these values are higher than our reported increased motion in AP and SI of the cerebral tonsils and brainstem. The discrepancies in the reported values could be due to the use of different methods to measure and compare the brain displacements. While in our work, we compared the 90^*th*^ percentile differences of the motion between the CM-I and healthy, in [29, 48] maximum displacements and peak-to-peak displacements have been compared. There are also differences between the imaging sequences, and other postprocessing steps. In [48], cine balanced fast-field echo MRI was used to scan the subjects, and the displacements were calculated manually. Whereas, in [29] DENSE MRI was used to scan the subjects, and an ROI was selected in the region with the maximum displacement. In our work, the displacement fields were calculated by the Demon’s image registration algorithm and 90^*th*^ percentile of the displacements were calculated in the masked brain structures.

In the next step, we analyzed the correlations between the morphometrics, brain displacement, and the surgical outcomes and symptomatology. Considering the limited number of CM-I subjects who received the decompression surgery (9 out 14), we only found the ML displacement of the tonsils to be predictive of the surgical outcomes (*p* = 0.03 and AUC = 0.95). For the symptomatology analysis, even though we did not find any statistical differences between the CM-I subjects with and without the symptoms, we observed a preliminary indication of 3D aMRI’s discriminative capability in these patients. In a male subject presenting with minor symptomatology (according to the supervising neurosurgeon), we observed the lowest peak cerebral tonsil displacement of about 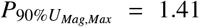 pixels amongst all the CM-I subjects. On the other hand, a male subject in his 20s with the most severe symptomatology had the second highest peak cerebral tonsil displacement of about 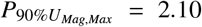 pixels. It should be noted that the average peak magnitude displacement amongst all the CM-I subjects was 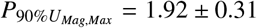 pixels.

These findings are of great importance, because even though the morphological parameters such as cerebral tonsil herniation are generally used for the diagnosis of CM-I, they often fail to predict the severity of the disease and the surgical outcomes [2, 6, 52]. Since a large number of CM-I patients have structural abnormality in the cerebellum but without underlying symptoms, morphological defects alone are not sufficient to cause symptomatic response. Therefore, dynamic information obtained from image processing methods such as 3D aMRI could provide us with rich information for better analysis of the underlying mechanisms of CMI. In this work, we have shown preliminary indications that the biomechanical information acquired from 3D aMRI can be used for symptomatology analysis and surgical decision making for the CM-I patients.

## Data Availability

All data produced in the present study are available upon reasonable request to the authors

## Acknowledgments

This research was supported by NIH Grant No. 1R21NS111415-01 and partially by NSF Grant No. CMMI-1953323.

